# Prevention of SARS-CoV-2 airborne transmission in a workplace based on CO2 sensor network

**DOI:** 10.1101/2022.03.04.22271934

**Authors:** Shinji Yokogawa, Yo Ishigaki, Hiroko Kitamura, Akira Saito, Yuto Kawauchi, Taisei Hiraide

## Abstract

We measured the compartmental air change per hour (ACH) using a CO_2_ sensor network in an office space where a cluster of COVID-19 infections attributed to aerosol transmission occurred. Generalized linear mixed models and dynamic time warping were used for a time series data analysis, and the results indicated that the ventilation conditions were poor at the time of the cluster outbreak, and that the low ACH in the room likely contributed to the outbreak. In addition, the adverse effects of inappropriate partitions and the effectiveness of ventilation improvements were investigated in detail. ACH of less than 2 /h was considered a main contributor for the formation of the COVID-19 cluster in the studied facility.

**Practical Implications:** A systematic method for measuring and evaluating indoor ventilation to prevent the spread of infectious diseases caused by aerosols is presented. Ventilation bias caused by ventilation pathways and inappropriate use of plastic sheeting can be detected by a CO_2_ sensor network and time series data analysis. Estimated ventilation rate will be a good index to suppress the formation of the COVID-19 cluster.

## Introduction

Controlling the spread of COVID-19 has become a priority worldwide. After case reports in December 2019, social distancing has been widely adopted as a containment strategy. The adoption of this social lifestyle has reaffirmed the importance of direct human connections and face-to-face interaction. Therefore, it is essential to control the risks and ensure safety in educational, public, and workplaces, which requires efforts of managers, supervisors, administrators, and all other stakeholders.

To reduce the risk of COVID-19 transmission, measures against all three routes of infection (contact, droplet, and aerosol) and to reduce the probability of infection through multiple defenses, social distance, mask, vaccine, etc., are required. However, compared to contact and droplet transmission, which can be prevented by social distancing and the use of masks, aerosol transmission is difficult to visually recognize, and the effectiveness of respective countermeasures has not been confirmed. Accordingly, mass transmissions of COVID-19 have been reported in poorly ventilated areas.^1^ In addition, the inappropriate use of plastic sheeting for preventing droplet infection has caused clusters of infectious diseases and threatened workplace safety.^2^ To avoid such risks, the use of CO_2_ sensors to control indoor air quality has attracted significant attention.^3-7^ The measurement of indoor CO_2_ concentration (referring to exhaled air) is considered an effective method for indirect risk management to ensure that exhaled aerosol particles containing SARS-CoV-2 do not remain indoors. Therefore, these devices have been widely installed as a safety measure in places where people gather, such as restaurants, stores, classrooms, and offices. The guideline for its operation considers a provisional control value of 800-1000 ppm set by government agencies of each countries as the maximum CO_2_ concentration.^8^ Under pandemic conditions, the Centers for Disease Control and Prevention has indicated that the CO_2_ concentration should be maintained below 800 ppm,^9^ which has been considered a good safety indicator.

However, as CO_2_ concentration is not a direct risk indicator and there is no direct epidemiological relationship between CO_2_ concentration and COVID-19 transmission, the effectiveness of this method should be verified to define appropriate control measures so as to improve the transmission risk and the safety of workplaces. In the manufacturing industry, quality assurance focuses on processes to ensure quality, and the proper control of a process is an indicator of product quality.^10^ Similarly, the risk management of COVID-19 requires the management and control of the environment (i.e., ventilation), rather than of the CO_2_ concentration itself.

The ventilation in a building room can generally be calculated by dividing the building volume by the ventilation volume of the installed ventilation measures. However, years after the construction of a building, the layout of rooms might change or the performance of the ventilation system can be degraded, so the capacity of the ventilation system will not be maintained as calculated. To overcome this issue, the ventilation in a target room can be evaluated and quantitatively assessed based on the CO_2_ concentration behavior monitored by CO_2_ sensors. In addition, the ventilation performance is not uniform for floors with large areas and complex layouts.^2,11^ Therefore, a systematic evaluation should be conducted by analyzing the time series of data from sensors arranged in a network, and these results can be used to improve ventilation and reduce transmission risks.

In this study, we used a CO_2_ sensor network and conducted a tracer gas experiment to evaluate the complex ventilation conditions in a business site in Japan, where a cluster of COVID-19 infections occurred. In 2021, 14 infections occurred among 30 workers who spent a short time in a large work preparation room with an area of 880 m^2^ and ceiling height of 3 m. The room was divided by four partitions, and only a small number of entrances and exits served as ventilation routes. The distance between the workers was greater than 2 m. The risk of contact and droplet infection was small, and the possibility of aerosol infection was high. In this study, we evaluated the air change rate for each partition considering the same conditions under which the COVID-19 cluster occurred and improved conditions, in which ventilation routes were determined. Based on this analysis, we performed a quantitative risk assessment. The data from the sensor network were statistically analyzed using generalized linear mixed models (GLMMs) and dynamic time warping to verify the effect of ventilation improvement. Based on the results and on our previous investigations, we discussed the ventilation index that reduces the occurrence of COVID-19 clusters. The results suggest that the improvement in ACH is reasonable and consistent with the lack of subsequent reports of infections.

### Objectives

The objectives of this research were to 1) demonstrate a method for evaluating and determining the state of air quality management in an office with a complex geometry using a CO_2_ sensor network, and 2) verify the effectiveness of ventilation improvement measures.

## Materials and Methods

This study was approved (approval number 21005) by the Ethics Committee on Experiments on Human Subjects of the University of Electro-Communications, Chofu, Tokyo, Japan.

The workplace where the cluster of infections occurred is a room where approximately 30 workers stayed for approximately one hour in the morning and one hour in the afternoon to prepare for the next work process. The workplace volume is approximately 2,640 m^3^, and it is divided by four partitions with a height of 1.8 m (Figure 1), so there was no face-to-face interaction and the possibility of droplet infection was low. The distance between adjacent workers was approximately 2 m. Employees work alone and are not required to communicate with each other, so communication between workers was kept to a minimum. In the next work process, the employees work at individual workstations that are far apart, so there was no physical contact.

**Figure 1.**
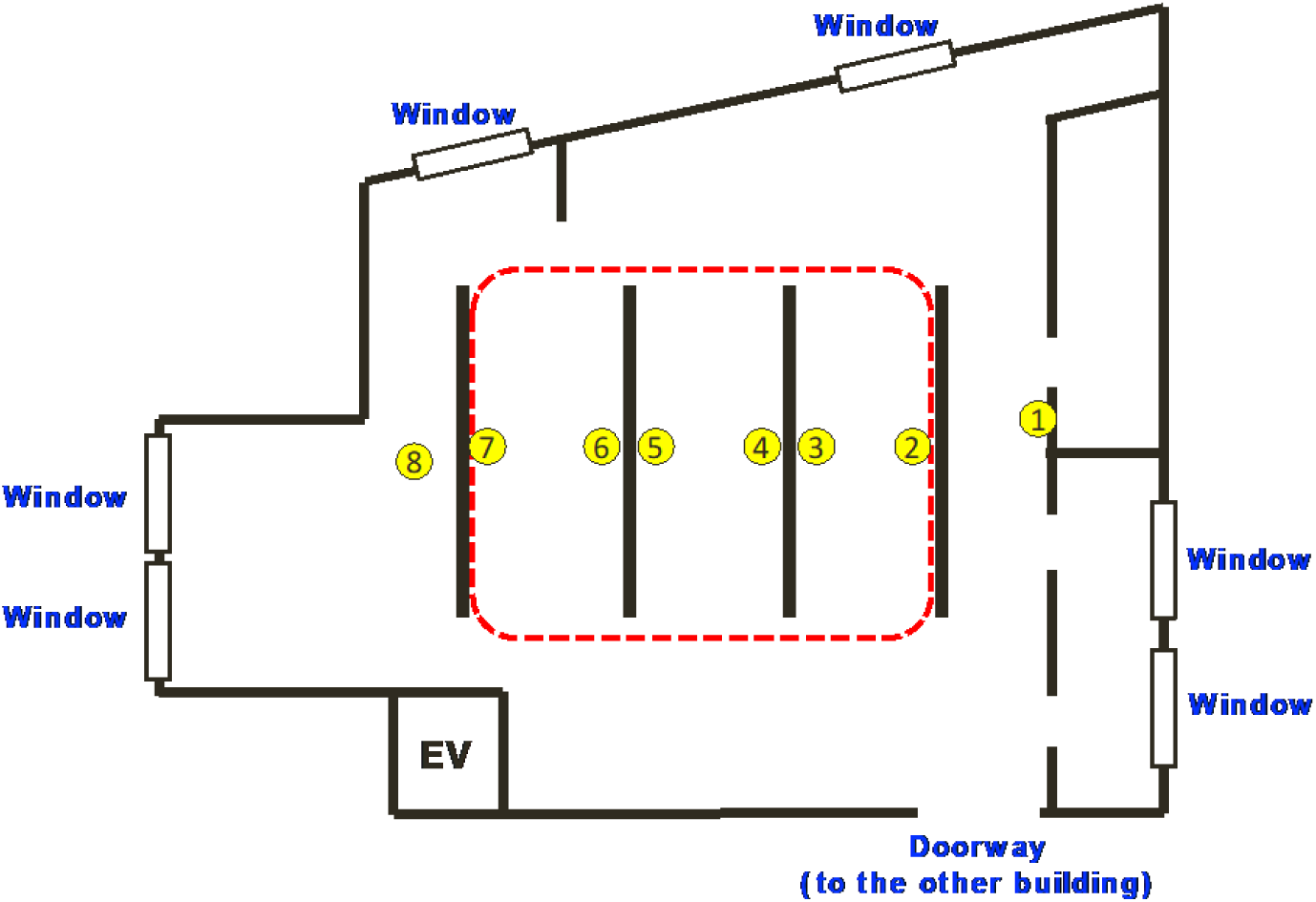
Layout of workplace and location of CO_2_ sensors.

The workplace is located on the second floor of a three-story building that was built in the 1950s and has been repeatedly remodeled. The main ventilation channels are the windows and doorways. During the cluster outbreak, although hand disinfection and masks were strictly enforced, only the doorway to another building was open for ventilation. This suggests that the risk of aerosol transmission was higher than that of contact or droplet transmission.

The COVID-19 cluster occurred in 2021. On weekdays, 52 employees work on this floor; 29 workers perform preparatory work for the next work step at the workbench in the red dotted frame in Figure 1. The other 23 managers and administrative staff work outside the red-dotted frame. One worker was the index case, and 10 of the 29 workers followed to be positive for COVID-19 according to the polymerase chain reaction (PCR) testing. In addition, one manager and one office worker also tested positive. The 29 workers stayed in the room at once for approximately one hour in the morning. In the afternoon, the number of people simultaneously in the room was less than 29 at all times. Employees only worked together for approximately one hour in the morning and in the afternoon they did not interact as a group, so the opportunity for mutual infection was limited. The relatively high infection rate among the workers, despite the short time spent in the compartment, suggests that the infection spread due to causes confined to the compartment. A test of the difference in infection rates using a normal approximation by logit transformation of the binomial distribution yielded a p-value of 0.02 for the null hypothesis of no difference in infection rates.

The compartmental indoor air ventilation was experimentally investigated. For that, the indoor airflow was observed using the CO_2_ tracer. Compartmental transmission risk was indirectly evaluated using the CO_2_ concentration as an alternative to the amount of exhaled air. In this study, dry ice was used as a CO_2_ source, and eight CO_2_ sensors were used to detect the changes in CO_2_ concentrations in each compartment, as shown in Figure 1.

Non-dispersive infrared (NDIR) gas sensors were used as the CO_2_ sensors. Eight TR-76Ui sensors (T&D Corporation, Japan) were placed on the desks within the compartments in which the employees had worked, as shown in Figure 1, in which the numbers 1 to 8 indicate the locations of each sensor. The TR-76Ui sensor can detect CO_2_ concentrations from 0 to 9,999 ppm, with an accuracy of ±50 ppm (±5%).

The experiment was conducted from 10:30 am to 12:30 pm on November 28, 2021. Dry ice was crushed on the floor to vaporize CO_2_, and the CO_2_ concentration in the room was increased to approximately 3,000 ppm, which is sufficiently larger than the background level of outdoor CO_2_, with no ventilation. After that, the decrease in CO_2_ concentration owing to the ventilation was measured by each sensor from 11:17 under ventilation Condition 1 (Table 1). After 35 min, the ventilation condition was changed to Condition 2 (Table 1), and the decrease in CO_2_ concentration attributed to ventilation was continuously measured from 11:52. Condition 1 reproduced the situation at the time of the cluster occurrence, and Condition 2 represented an improved ventilation condition.

**Table 1.** Ventilation conditions of experiment.

|  | Condition 1 | Condition 2 |
| --- | --- | --- |
| Time | 11:17 to 11:52 | 11:52 to 12:08 |
| Doorway | Open | Open |
| Window | Closed | Open |

The air change per hour (ACH), which represents the ventilation around sensors, was estimated based on the time series change in CO_2_ concentration under Condition 1. A transient mass balance model was used to solve the CO_2_ concentration around the sensors. The stable mass balance of well-mixed air can be described as:

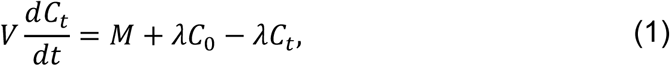

where *C*_*t*_ is the concentration of indoor pollutants at time *t, M* is the number of pollutants generated, *λ* is the air ventilation rate, and *C*_0_ is the concentration in the absence of pollutant sources. As a general solution to the first-order linear ordinary differential equation shown in Equation (1), the concentration of pollutants at time *i* can be obtained by the Seidel’s equation^12,13^:

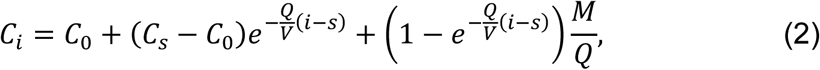

where *Q* is the outdoor air supply around the sensors [m^3^/h], *V* is the effective volume of the space around the sensors, *s* is the time at which the observation started, and *λ = Q*/*V* is the assumed air ventilation rate. When no pollutants are generated, i.e. *M =* 0, Equation (2) can be transformed into:

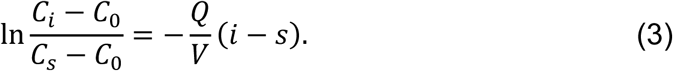

This equation suggests that a decrease in the normalized concentration of CO_2_ with the ventilation time (*i* − *s*) [h] without pollutant generation corresponds to the ACH in the space, which was estimated using the linear model of *Q*/*V* [/h] from Equation (3).

In this study, a 95% confidence interval for the estimated ACH was calculated along with the point estimates. The GLMM was used to statistically verify the improvement in ACH and its dependence on the position relative to ventilation points.

## Results

The observed CO_2_ behavior reflected well the characteristics of the workplace compartmental ventilation. Figure 2 shows a time series variation of the observed CO_2_ concentration over the experimental period. After increasing the indoor CO_2_ concentration from 2500 to 3000 ppm, which are significantly higher than the outdoor background concentration, the windows and entrances were set to ventilation Condition 1, and a gradual decrease in CO_2_ concentration was observed in all locations of the room. Even after 34 min of ventilation, the CO_2_ concentration only dropped to approximately 1500–2250 ppm. In addition, the variability in the CO_2_ concentration also increased compared to that in the beginning of the observation period. Therefore, we changed the settings of the windows and entrances to ventilation Condition 2 to create a ventilation path, and observed the changes in CO_2_ concentration for 16 min. A slight improvement in the ventilation rate was observed. In addition, the variability in the CO_2_ concentration decreased slightly.

**Figure 2.**
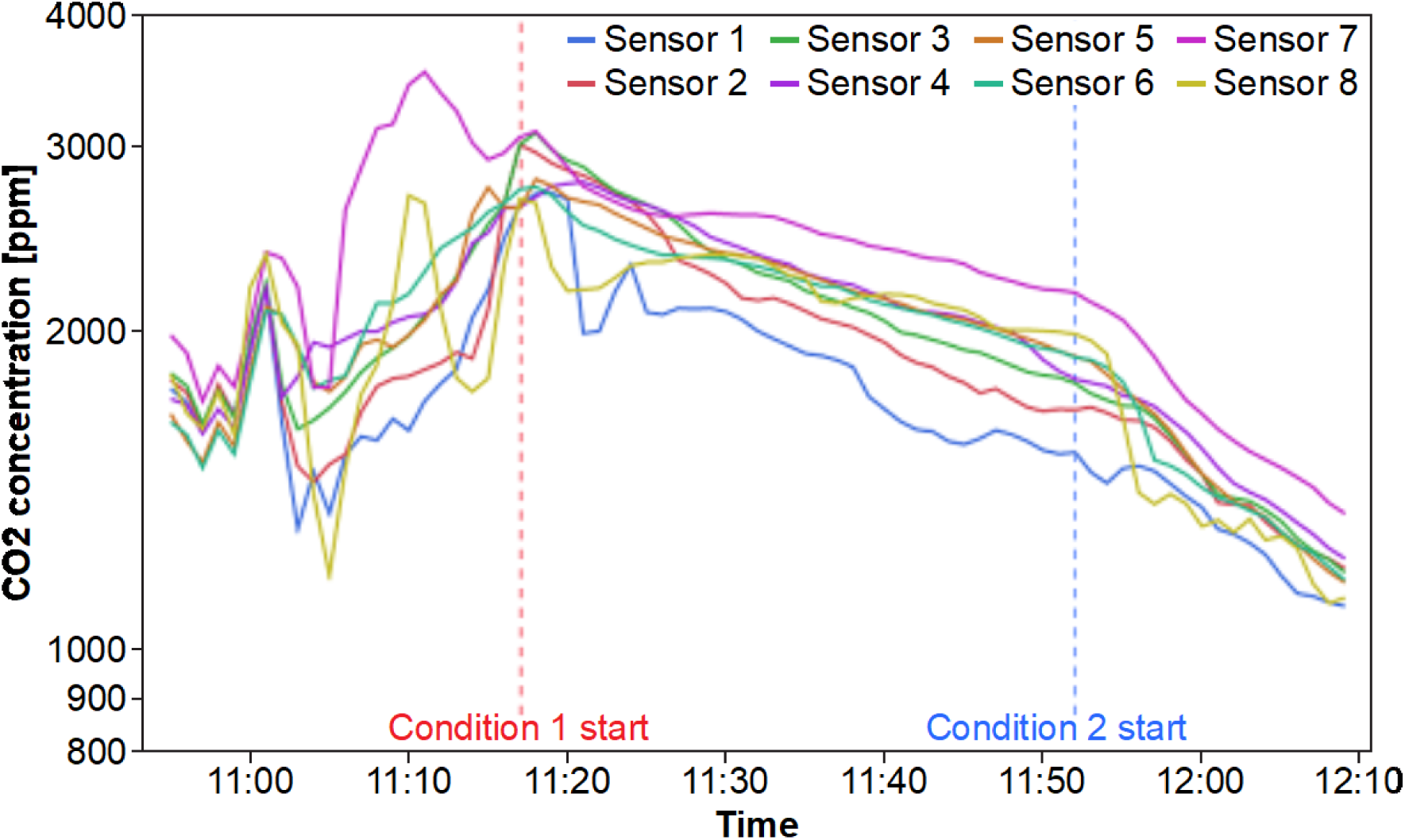
CO_2_ concentration observed during experiments.

The changes in CO_2_ concentration observed by the implemented experimental method fitted well Equation 2, so we can conclude that ACH was estimated with high accuracy. The normalized difference between the observed CO_2_ concentration and the background concentration (assumed as 400 ppm) was plotted against the time of observation, as shown in Figure 3. There was a clear difference in the slope of the fitted line between ventilation Conditions 1 and 2. Under Condition 1, the plot was divided into two groups: sensors 1–3 and 4–8. This was attributed to the fluctuations immediately after the start of the observation periods. Therefore, it is reasonable to focus on the slope, that is, the number of compartmental ventilations, to quantify the compartmental ventilation.

**Figure 3.**
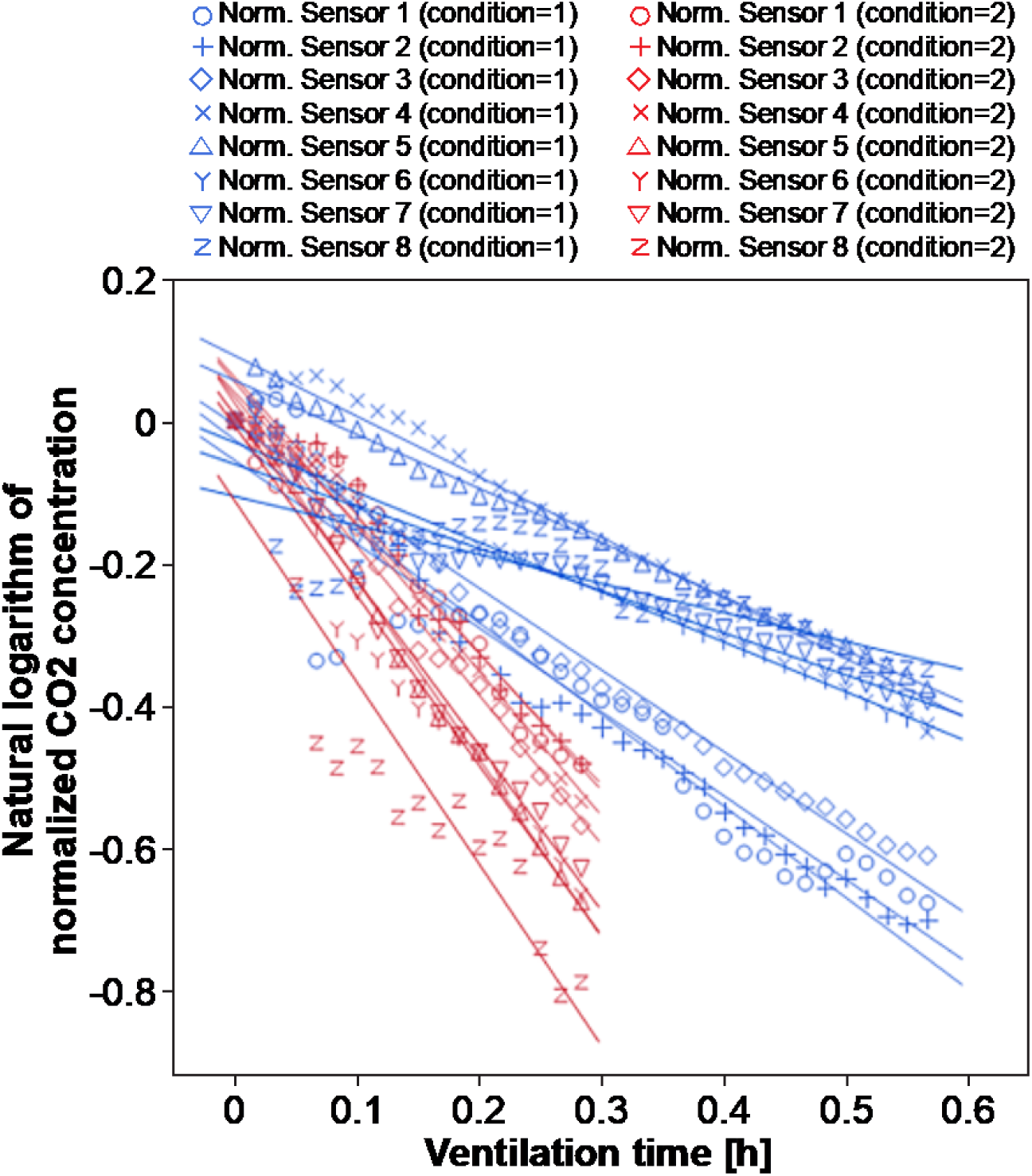
Analysis of compartmental air change per hour (ACH). Slopes of fitted lines indicate ACH around sensors.

The ACH was estimated from the slope of the straight line in Figure 3. Although Equation (2) is linear and without an intercept, the intercepts for each line cannot be neglected because of the fluctuations in the first observations. Therefore, to ensure estimation accuracy, the slope was estimated by assuming a linear equation with an intercept. The ACHs estimated for each condition and sensor are shown in Table 2. The statistical software JMP Pro Ver. 16 was used for the regression analysis.

**Table 2.**
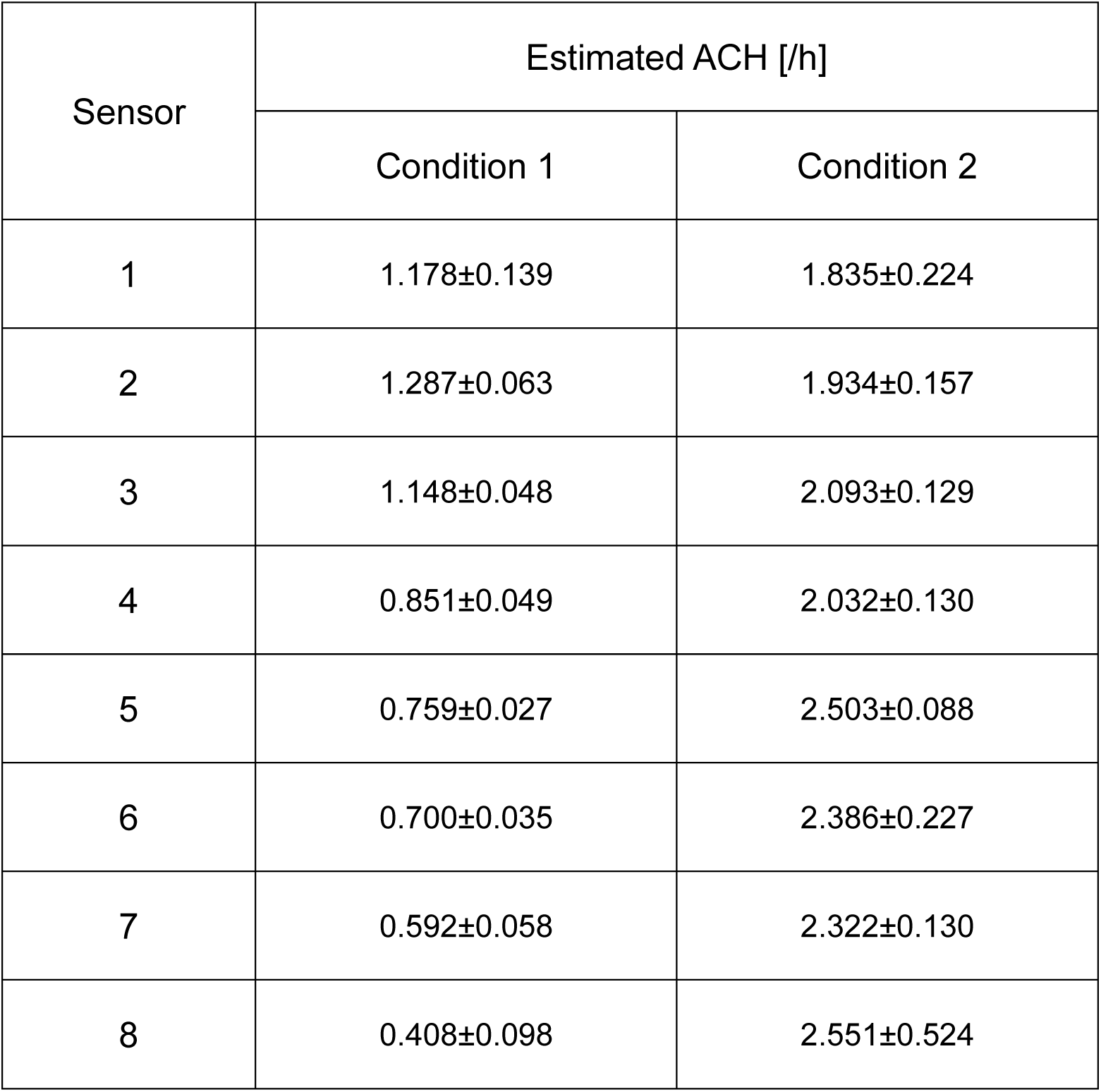
Estimated air change per hour (ACH) (95% confidence interval) for each condition and sensor based on a linear regression analysis.

Figure 4 shows the estimated ACH values. Under Condition 1 (the same during the COVID-19 cluster outbreak), the ACH was low, and its maximum value was less than 1.5 /h. Moreover, the ACH decreased as the distance from the doorway increased because the doorway was the only route for the introduction of outside air. At the farthest point, near sensor 8, ACH was less than 0.5 /h. According to the Japanese Ministry of Health, Labor and Welfare (MHLW), in workplaces without proper ventilation structures, ventilation should be provided at least twice an hour by opening windows and doorways.^8^ Furthermore, the probability of tuberculosis infection, for which airborne transmission is the established route of infection, is markedly reduced in workplaces with an ACH of 2 or higher.^14-16^ The ACH at the time of the COVID-19 cluster outbreak was probably much lower than this.

**Figure 4.**
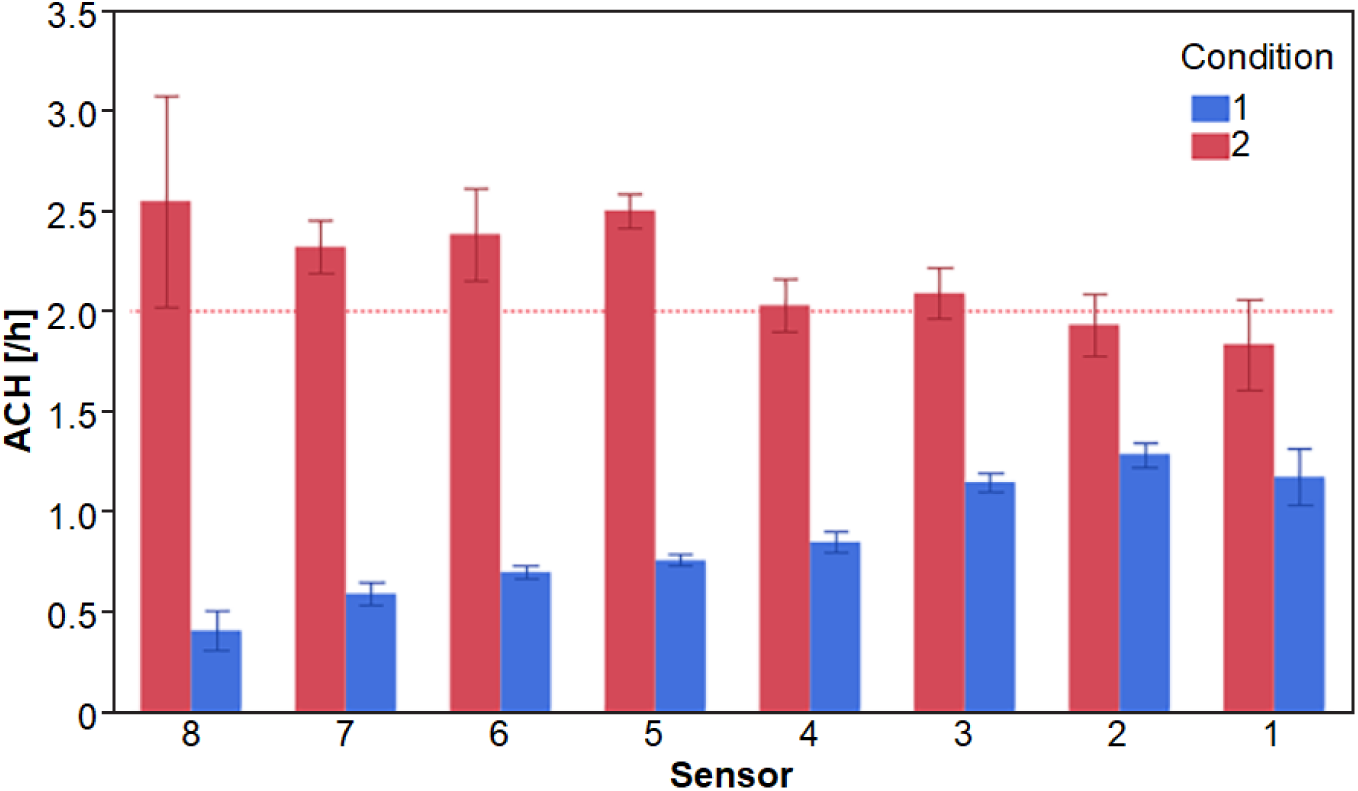
Estimated compartmental air change per hour (ACH) according to sensor. Error bars indicate a 95% confidence interval.

Under Condition 2, in which a window is opened to form a ventilation path, ACH was improved to approximately 2 /h. In addition, the ACH trend by location was opposite to that under ventilation condition 1; its value increased as the distances from the doorway increased. That is, the amount of outside air introduced by opening the window was dominant under ventilation condition 2.

## Discussion

In the Results section, we analyzed the data from each individual sensor and compared the results to identify the respective trends. However, it is also necessary to statistically test and estimate the trend of ACH variation based on time series data analysis to clarify if the variation was significant compared to the observation errors.

The GLMM^17^ was used to analyze the effects of experimental conditions on ventilation and the differences and trends in ACH attributed to sensor location, that is, the inhibitory effects of inappropriate partitions on ventilation. The GLMM model is a mixed effect model in which the ventilation time and interaction between ventilation time and sensor location are fixed effects, and the sensor location is a random effect. The objective variable is the ratio of the increase in CO_2_ concentration from the background to that at the beginning of the observation, and the model presented a natural logarithm as the link function.

The GLMM analysis was conducted using the statistical software JMP Pro Ver. 16 for Conditions 1 and 2. In both cases, the GLMM explained the observed data well, with a coefficient of determination R2 > 0.96. The results of the fixed-effects tests are listed in Table 3. The estimated covariance parameters of the variational effects are shown in Table 4. The results shown in Table 3 indicate that the interactions between ventilation time and sensor location were highly significant. Moreover, the decrease in CO_2_ concentration with ventilation time for Conditions 1 and 2 was also highly significant. The results shown in Table 4 also indicate that the random effects were not significant on their own for both Conditions 1 and 2. These results suggest that ventilation was impeded by the partitions installed in the room, and that the ACH fluctuation depends on the position in the partitions and air flow root.

**Table 3.**
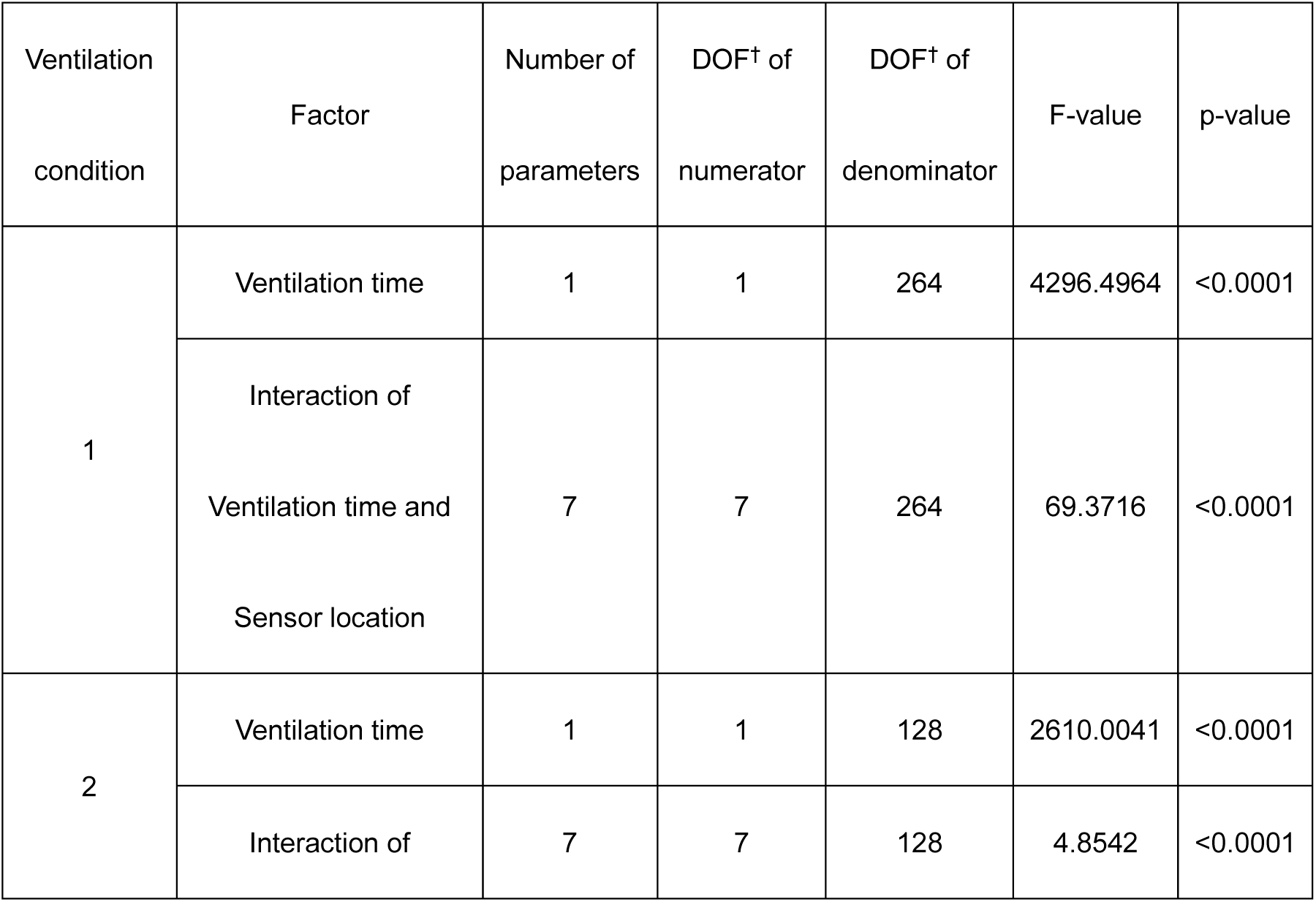
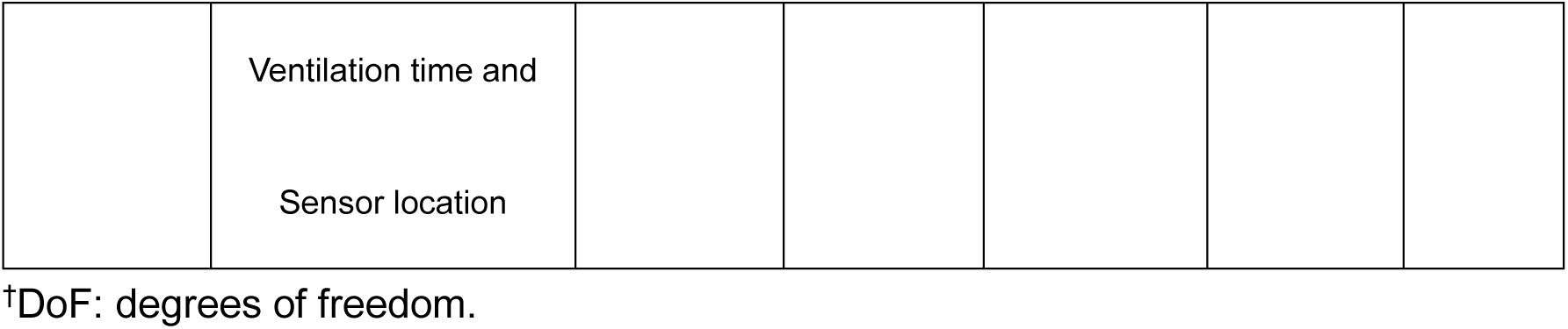
Results of the test of fixed effects for each ventilation condition.

**Table 4.** Covariance parameter estimates of random effects according to ventilation condition.

| Ventilation condition | Variance components | Estimate | Standard error | 95% Lower limit | 95% Upper limit | Wald p-value |
| --- | --- | --- | --- | --- | --- | --- |
| 1 | Sensor | 0.0092261 | 0.0049527 | -0.000481 | 0.0189332 | 0.0625 |
|  | Residual error | 0.0013827 | 0.0001204 | 0.0011742 | 0.0016526 |  |
|  | Sum | 0.0106089 | 0.0049541 | 0.0050479 | 0.0348439 |  |
| 2 | Sensor | 0.0075253 | 0.0040821 | -0.000476 | 0.0155261 | 0.0653 |
|  | Residual error | 0.0020094 | 0.0002512 | 0.0015955 | 0.0026093 |  |
|  | Sum | 0.0095347 | 0.004089 | 0.0047692 | 0.0276991 |  |

The maximum likelihood estimates of ACH for each condition and sensor are listed in Table 5. They were in good agreement with the results of the individual linear regressions, thereby supporting the validity of the discussion obtained from Tables 3 and 4.

**Table 5.** Air change per hour (ACH) (95% confidence level) according to ventilation conditions and sensors estimated by a generalized linear mixed model.

| Sensor | Estimated ACH [/h] |  |
| --- | --- | --- |
|  | Condition 1 | Condition 2 |
| 1 | 1.178±0.073 | 1.835±0.239 |
| 2 | 1.287±0.073 | 1.934±0.239 |
| 3 | 1.148±0.073 | 2.093±0.239 |
| 4 | 0.851±0.073 | 2.032±0.239 |
| 5 | 0.759±0.073 | 2.503±0.239 |
| 6 | 0.701±0.073 | 2.386±0.239 |
| 7 | 0.592±0.073 | 2.322±0.239 |
| 8 | 0.408±0.073 | 2.551±0.239 |

Figure 5 shows the fitting results of the estimated GLMM to the observed values. The estimated GLMMs explain the observed values well, which indicates that the ACH estimates and their variations were reasonable. However, the observed values for sensors 1 and 8, which were located at the outermost periphery of the room, showed relatively large errors compared to the other sensors. This might have occurred because the amount of ventilation introduced from outside air was affected by the weather, wind speed, and wind direction outside the room. Conversely, the ventilation in the inner partitions was less sensitive to the outdoor environment.

**Figure 5.**
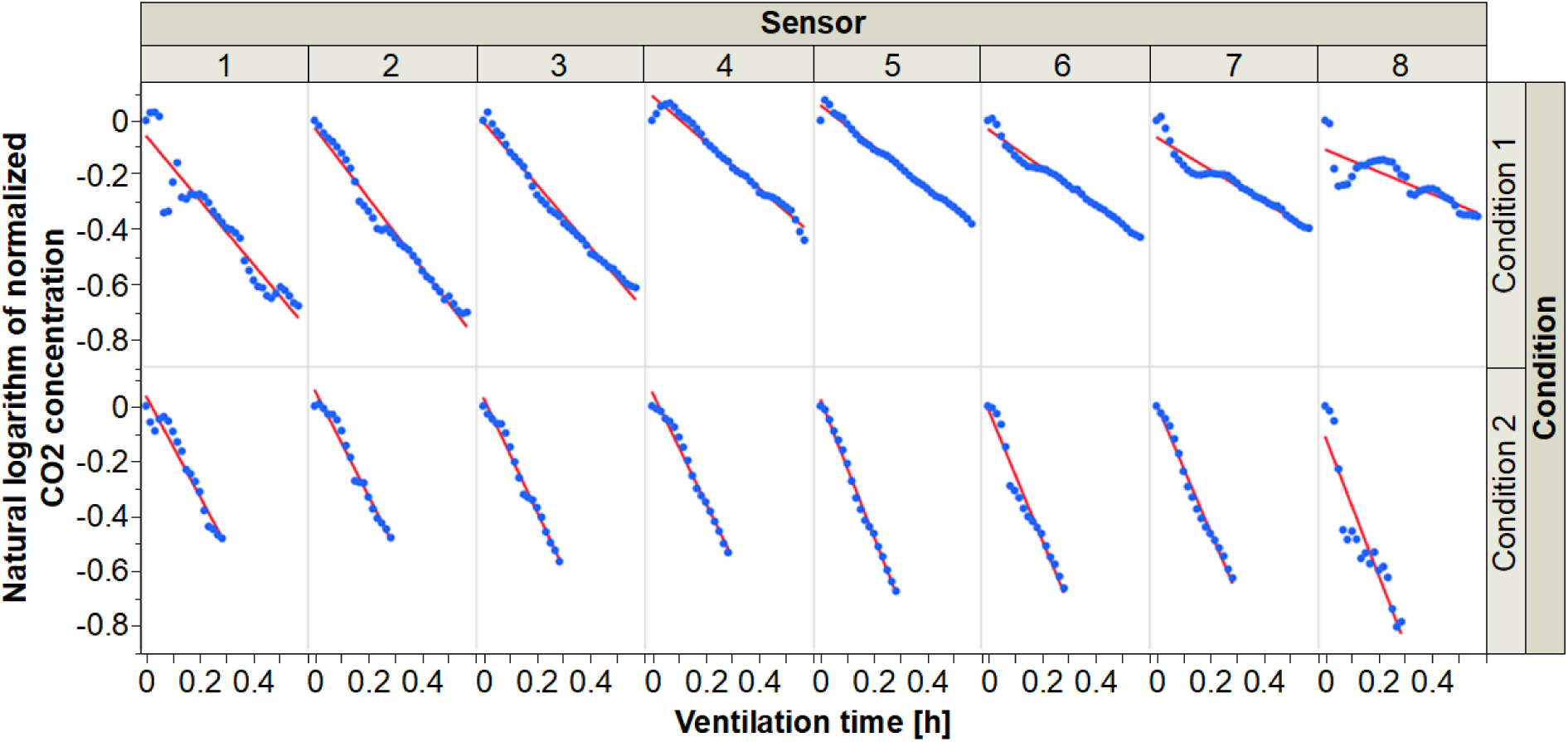
Fittings of estimated generalized linear mixed models to observed CO_2_ concentrations.

The characteristics of the ventilation distribution were investigated based on the similarity of the ACH patterns at each sensor location. The dynamic time warping (DTW)^18,19^ method was used to calculate the DTW distance of the ventilation patterns measured over time. DTW is an algorithm for measuring the similarity between two time-series data, which may vary in speed. Similarities in CO_2_ variation can be detected using DTW, even if there are accelerations and decelerations during an observation. To calculate the DTW distance, we used the statistical language R package “dtw” Ver. 1.22-3.

The matrix of the DTW distance between each sensor data is shown in Figure 6. The DTW matrices were calculated for each ventilation condition. They were colored according to the distance, with a relatively high pattern similarity colored in green and low pattern similarity colored in red in a stepwise manner. The diagonal component was excluded because it refers to itself. Because of the difference in the number of data, the absolute values of the DTW distance between Conditions 1 and 2 could not be compared. Therefore, the study focused on the difference in room similarity distribution.

**Figure 6.**
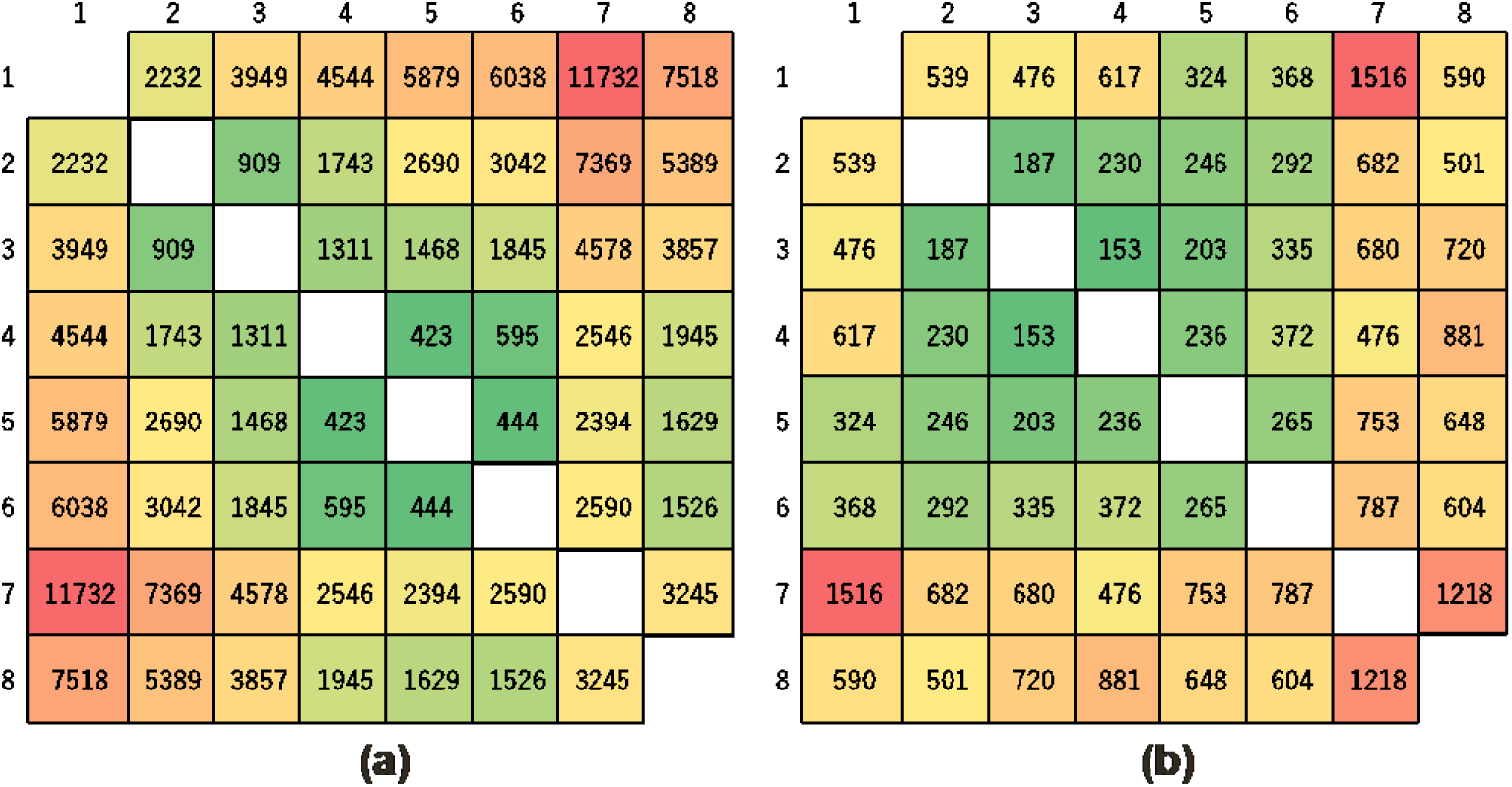
DTW distance matrices for each ventilation condition. (a) Ventilation condition 1. (b) Ventilation condition 2.

Figure 6 shows that sensors 1, 7, and 8, which are near the edge of the room, presented less similarity in behavior with the sensors near the center of the room. The results suggest that the ventilation patterns of neighboring locations were similar. The range of pattern similarity near the center of the room was wider under Condition 2 than under Condition 1. These results suggest that although opening the windows in ventilation Condition 2 improved the ventilation of the entire workplace, the ventilation effect was less effective in reaching the center of the room because of the partitions. Therefore, it was speculated that ventilation can be increased by improving the airflow along the partitioned areas.

In summary, for aerosol infection control, which should be conducted in parallel with measures against contact and droplet transmission, the maximum height of partitions should be strictly controlled, and they should be installed at a height and orientation that do not interfere with ventilation. Measurement using sensor networks is effective in detecting such a ventilation bias. In addition, the observed bias of CO_2_ is more complex in rooms with larger sizes, complex geometries, and various uses,^2,11^ and this study agrees with previously reported results.

Although there is still a possibility of confounding variables owing to the relatively small number of infections in Japan since October 2021, the fact that no infections have been confirmed since the adoption of improved ventilation Condition 2 and the results of our previous study^2^ indicate that the outbreaks of the infection clusters share the feature of a ACH of less than 2 /h. In addition, where the ACH was improved to 2 or more /h, no evidence of a second infection cluster has been identified.

This index for ACH is consistent with the results of previous studies on tuberculosis and is considered highly valid.^14-16^ In large workplaces with complex layouts and partitions, ventilation conditions become complex, so local monitoring and quantitative evaluation using sensor networks, as shown in this study, are effective.

In future studies, real-time data from the CO_2_ sensor network should be analyzed to identify compartments with increased risk to issue alerts at appropriate times. This would require the combination of CO_2_ concentration and other sensor data such as temperature, humidity, illumination, barometric pressure, and human detection. The authors have already reported a method of applying topological data analysis to multidimensional time-series data from many sensors.^11^ In the future, we hope to develop a method for the diagnosing of anomalies by combining data integration with machine learning and deep learning.

## Conclusions

We measured ACH using a CO_2_ sensor network in a workplace where a cluster of COVID-19 infections occurred, and investigated the adverse effects of inappropriate partitions and the details of the ventilation improvement effect. A statistical analysis using GLMM showed that the ACH in the room was biased along to the position in the partitions and air flow root. In addition, by examining the similarity of ventilation patterns using DTW, the distribution of ventilation patterns between the partition and the effect of ventilation conditions were evaluated. The results indicated that a ACH of less than 2 /h was favorable for the formation of theCOVID-10 clusters in this facility.

## Data Availability

All data produced in the present study are available upon reasonable request to the authors

## Acknowledgments

This work was supported by JSPS KAKENHI Grant No. 21K19820.

